# Molecular pathology of acute respiratory distress syndrome, mechanical ventilation and abnormal coagulation in severe COVID-19

**DOI:** 10.1101/2023.03.09.23286797

**Authors:** Antoine Soulé, William Ma, Katelyn Yixiu Liu, Catherine Allard, Salman Qureshi, Karine Tremblay, Amin Emad, Simon Rousseau

## Abstract

Systemic inflammation in critically ill patients can lead to serious consequences such as acute respiratory distress syndrome (ARDS), a condition characterized by the presence of lung inflammation, edema, and impaired gas exchange, associated with poor survival. Understanding molecular pathobiology is essential to improve critical care of these patients. To this end, we use multimodal profiles of SARS-CoV-2 infected hospitalized participants to the Biobanque Québécoise de la COVID-19 (BQC-19) to characterize endophenotypes associated with different degrees of disease severity. Proteomic, metabolomic, and genomic characterization supported a role for neutrophil-associated procoagulant activity in severe COVID-19 ARDS that is inversely correlated with sphinghosine-1 phosphate plasma levels. Fibroblast Growth Factor Receptor (FGFR) and SH2-containing transforming protein 4 (SHC4) signaling were identified as molecular features associated with endophenotype 6 (EP6). Mechanical ventilation in EP6 was associated with alterations in lipoprotein metabolism. These findings help define the molecular mechanisms related to specific severe outcomes, that can be used to identify early unfavorable clinical trajectories and treatable traits to improve the survival of critically ill patients.

## Introduction

Acute respiratory distress syndrome (ARDS), a condition characterized by the presence of lung inflammation, edema, and impaired gas exchange, is observed in severe coronavirus disease 2019 (COVID-19) and associated with poor survival. Mechanical ventilation is an important critical care management strategy for severe COVID-19 (1). However, during the first waves of the pandemic, almost half of patients undergoing invasive mechanical ventilation died based on case fatality rate reports (2). Defining the molecular mechanisms related to specific severe outcomes, such as ARDS or acute respiratory failure that require invasive mechanical ventilation, is important to identify treatable traits and improve the survival of critically ill patients.

Endophenotypes are subgroups which are inapparent to traditional methods but share a common set of factors that dictate their response in a manner that is distinct from other sub-groups (3). Endophenotypes have been described in airway diseases such as asthma (4), COPD, (5) and chronic rhinosinusitis (6). Recent studies have identified endophenotypes associated with COVID-19 severity using different large data sets, such as bulk transcriptomic from blood (7), bulk transcriptomic from neutrophil-enriched fractions (8), or proteomic in our accompanying manuscript (9).

In this manuscript, we report multimodal characterization of one of the six endophenotypes (EPs) identified in our accompanying manuscript (9), which we found to be associated with the worst clinical trajectory during hospitalization following SARS-CoV-2 infection (including ARDS). This analysis reveals known and novel molecular effectors of immuno-thrombosis, ARDS and mechanical ventilation associated with COVID-19.

## Results

### Serprocidins are enriched in an endophenotype associated with COVID-19 severity and ARDS

To gain insight into the molecular mechanisms underlying the severity in COVID-19, we investigated the aptamers enriched within an endophenotype (EP6) (Figure 1 and Table S1) that we previously discovered using the circulating proteome of SARS-CoV-2 infected and hospitalized participants to BQC19 (n = 731) and found to be associated with severe outcome and ARDS (9). These aptamers correspond to measurements obtained using a multiplex SOMAmer affinity array. We used two-sided Mann–Whitney U (MWU) tests and the p-values were corrected for the number of aptamers and EPs using Benjamini–Hochberg (BH) false discovery rate (FDR). Consistent with the elevated neutrophil numbers in EP6, the most enriched aptamer was Cathepsin G (FDR = 4.10E-58), a serine protease localized in neutrophil azurophil granules. Notably, two other serprocidins, neutrophil elastase and proteinase-3, were also significantly enriched in EP6 (FDR = 1.04E-10 and FDR = 8.27E-7, respectively, Figure 1 and Table S1). Moreover, another NETosis-associated protein, Olfactomedin 4 (OLFM4) (10), was greatly enriched in EP6 (top 5%; FDR = 2.61E-46, Figure 1 and Table S1). This suggests an important role for neutrophil degranulation and/or NETosis in severe COVID-19.

**Figure 1:**
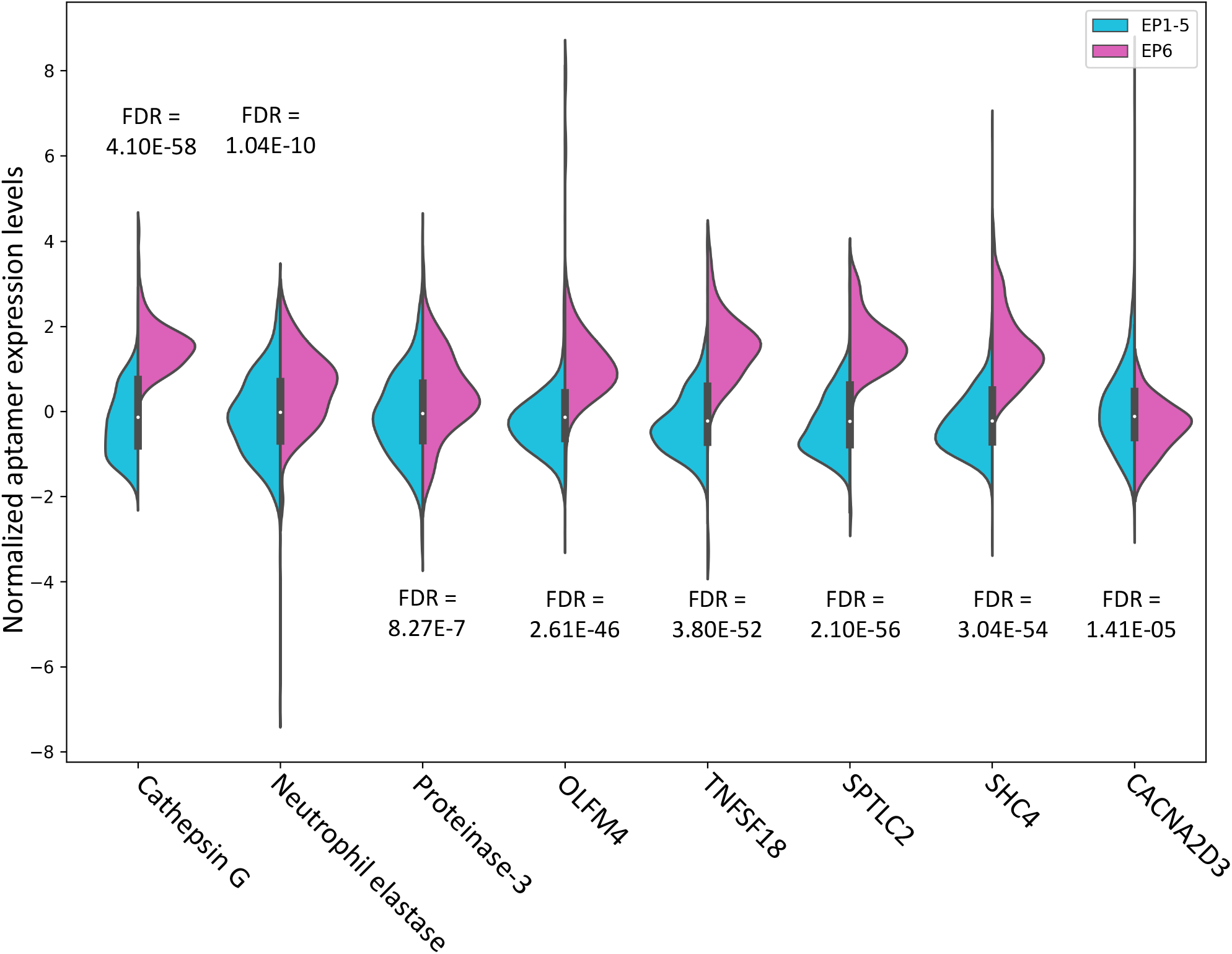
Distribution of aptamer expressions in EP6 versus other EPs. The violin plots show the distribution of aptamer expressions for patients in EP6 (magenta) versus EP1-5 (teal). Two-sided Mann–Whitney U (MWU) tests followed by Benjamini–Hochberg method for adjusting the p-values were used to calculate false discovery rates.

### Pathway enrichment analysis identified FGFR-signaling in severe COVID-19 acute respiratory distress syndrome

To capture a more comprehensive landscape of the signaling networks active in EP6, we performed pathway enrichment analysis using the Knowledge Engine for Genomics (KnowEnG) (11). Since aptamers were used to identify the EPs, it is expected that many of them would have significant associations with the EPs. As a result, we selected a very strict threshold of FDR < 1E-20 (two-sided MWU test, BH FDR) to identify top aptamers associated with EP6 (9). We then used the gene set characterization pipeline of KnowEnG with Reactome pathway collection (12). This analysis showed that EP6 is characterized by pathways associated with interleukins and cytokine signaling in the immune system. In addition, fibroblast growth factor receptor (FGFR) signaling was identified in EP6, suggesting this pathway as a potential driver of severe pathology (Table 1 and S1).

**Table 1:**
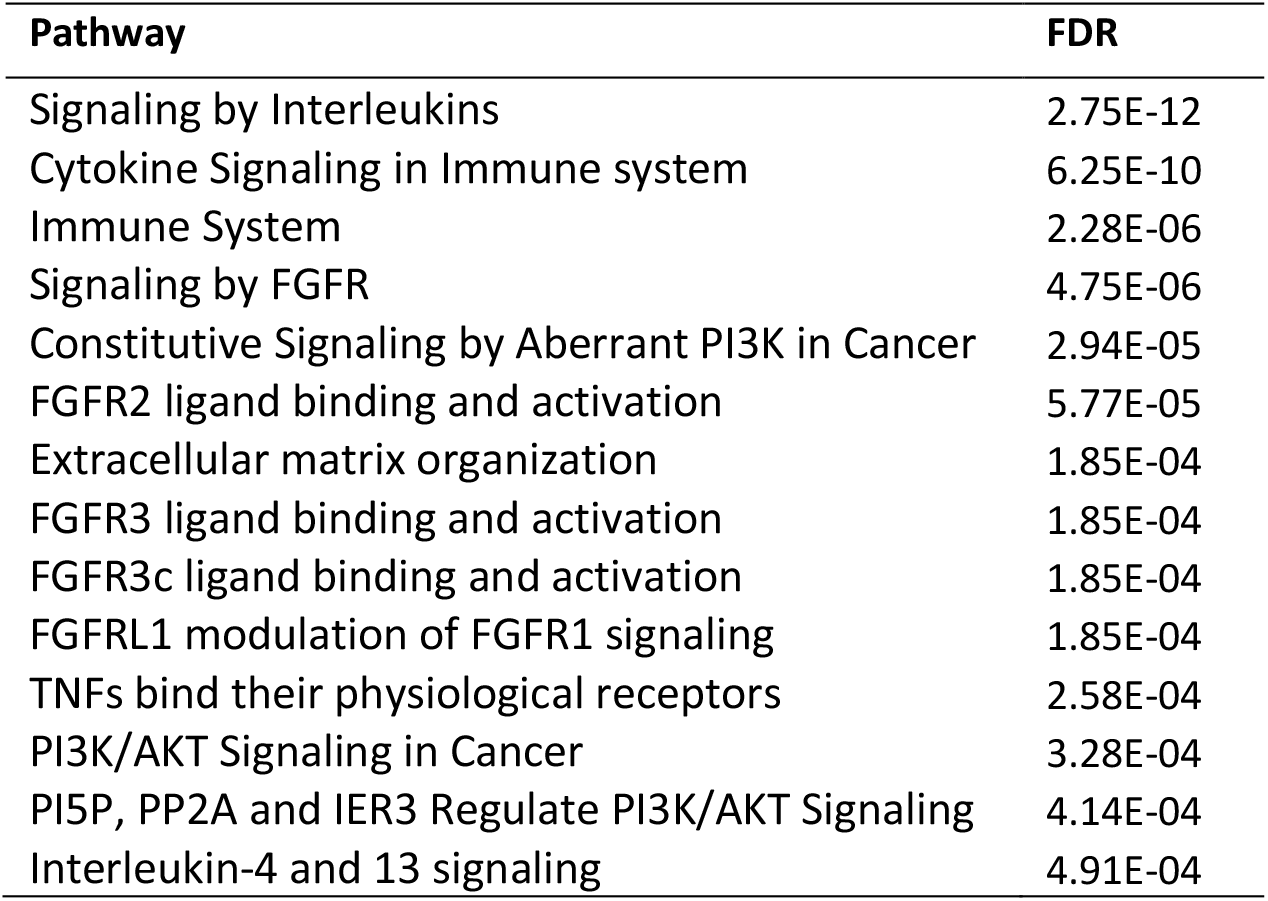
Reactome pathways associated with EP6, based on expression of aptamers. KnowEnG analytical platform was used. The p-values were calculated using a one-sided Fisher’s exact test and were corrected for multiple tests using Benjamini–Hochberg method. Only signaling pathways with FDR < 5E-4 are shown in this table (see Table S1 for the full list).

### EP6 metabolomic signature was associated with pro-coagulant activities and liver dysfunction

To further characterize the pathobiology of severe COVID-19 disease, metabolomic profiling of plasma samples was performed in parallel to the aptamer analysis. The results yielded data on 1,435 metabolites, of which 576 were found significantly altered in EP6 (two-sided MWU FDR < 0.01) (Table S2). Characterization of the plasma samples supported the distinction among the different EPs at the levels of metabolomic sub-pathways and individual metabolites (Figure 2 and Table S2). The two top sub-pathways (FDR < 0.01) “Methionine, Cysteine, SAM and Taurine Metabolism” and “phosphatidinylethanolamine (PE)”, are known to interact (13, 14) and are associated with pro-coagulant activities. Phospholipids-containing microparticles from platelet activation contribute to Tissue Factor activation and pro-thrombinase activity (15), linking PE to coagulation and D-dimers level, which are elevated in EP6 (9). PE methylation acts as a major consumer of S-Adenosylmethionine (SAM) leading to the synthesis of S-Adenosylhomocysteine (SAH) and cystathionine (14), upstream of 2-hydroxybutyrate and 2-aminobutyrate in a model of hepatoxicity (16). SAH, cystathionine, 2-hydroxybutyrate and 2-aminobutyrate are all significantly enriched in EP6 (Figure 2B and 3A). To identify relationships that may shed light on factors influencing clinical laboratory results that define EP6, we also performed Spearman’s rank correlation analyses between 21 blood variables and metabolites (Table S3). SAH is positively correlated with creatinine, while cystathionine is positively correlated with creatinine and negatively correlated with albumin (Figures 4, S1-S2), congruent with the data from the model of hepatoxicity (16), linking abnormal coagulation and liver damage in severe COVID-19. We also observed kynurenine to be enriched in EP6 but depleted in EP1 (Figures 3 and 5). Kynurenine levels were also found to be positively correlated with D-Dimers levels (Figure 4 and S1-S2), suggesting a role of this metabolite of the tryptophan pathway in abnormal coagulation in severe COVID-19. Interestingly, TNFSF18 (GITR), a T-cell receptor that can activate indoleamine 2,3-dioxygenase (IDO) to increase kynurenine synthesis (17) in allergy, is enriched in EP6 and depleted in EP1 (Figures 3 and 5).

**Figure 2:**
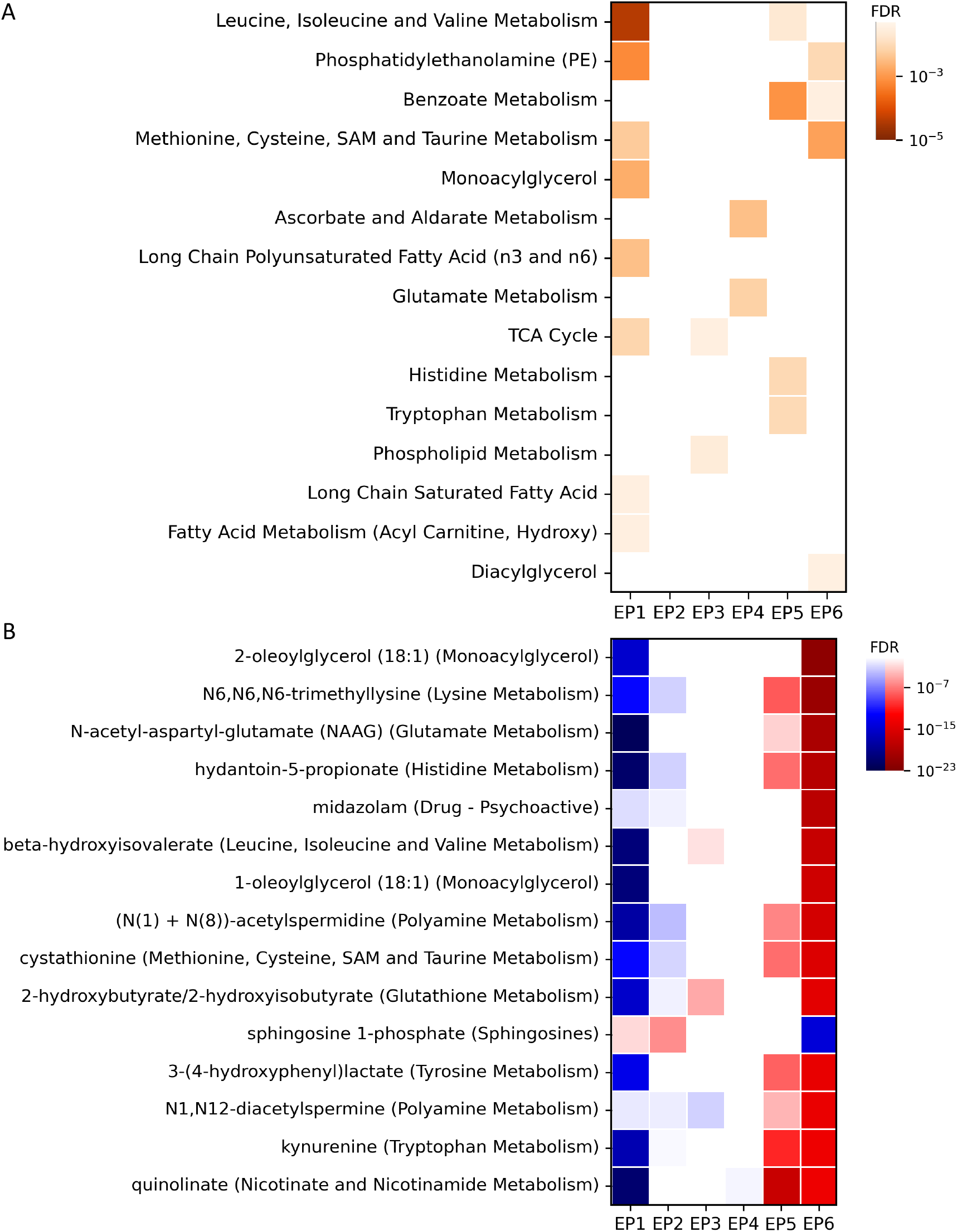
Metabolite characteristics of endophenotypes (EPs). A) The heatmap shows the enrichment (one-sided Fisher’s exact test) of EPs in different metabolite sub-pathways. See Table S2 for the full list. B) The heatmap shows the over-expression (red) and under-expression (blue) of metabolites in different EPs (two-sided Mann– Whitney U test). Row names show metabolites followed by the sub-pathway to which they belong in parentheses. Only top 15 metabolites (based on false discovery rate for EP6) for which a definite name and sub-pathway was available are shown. Full list is provided in Table S2.

**Figure 3:**
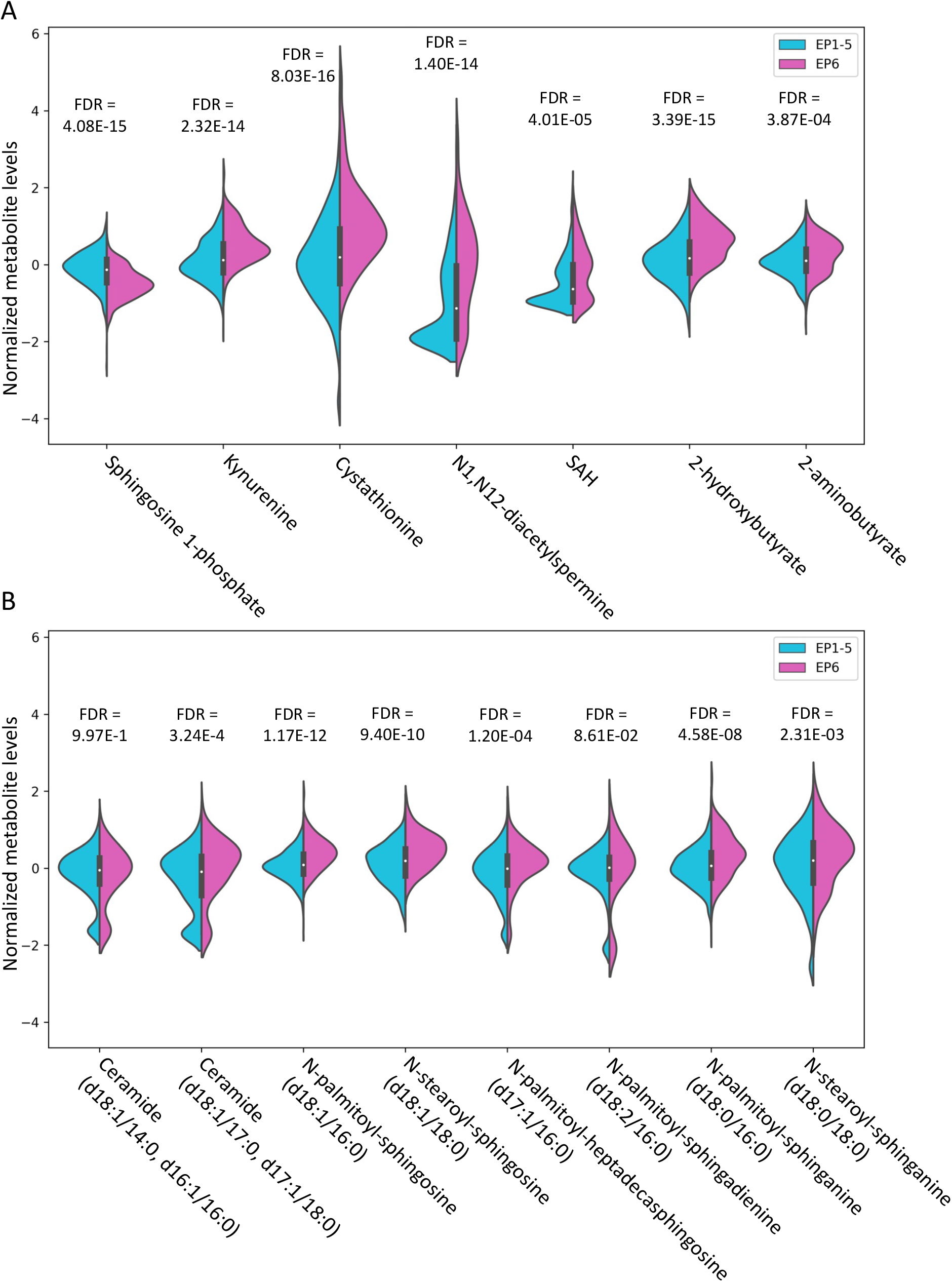
Distribution of metabolite levels in EP6 versus other EPs. The violin plots show the distribution of metabolite levels for patients in EP6 (magenta) versus EP1-5 (teal). Two-sided Mann–Whitney U (MWU) tests followed by Benjamini–Hochberg method for adjusting the p-values were used to calculate false discovery rates.

**Figure 4:**
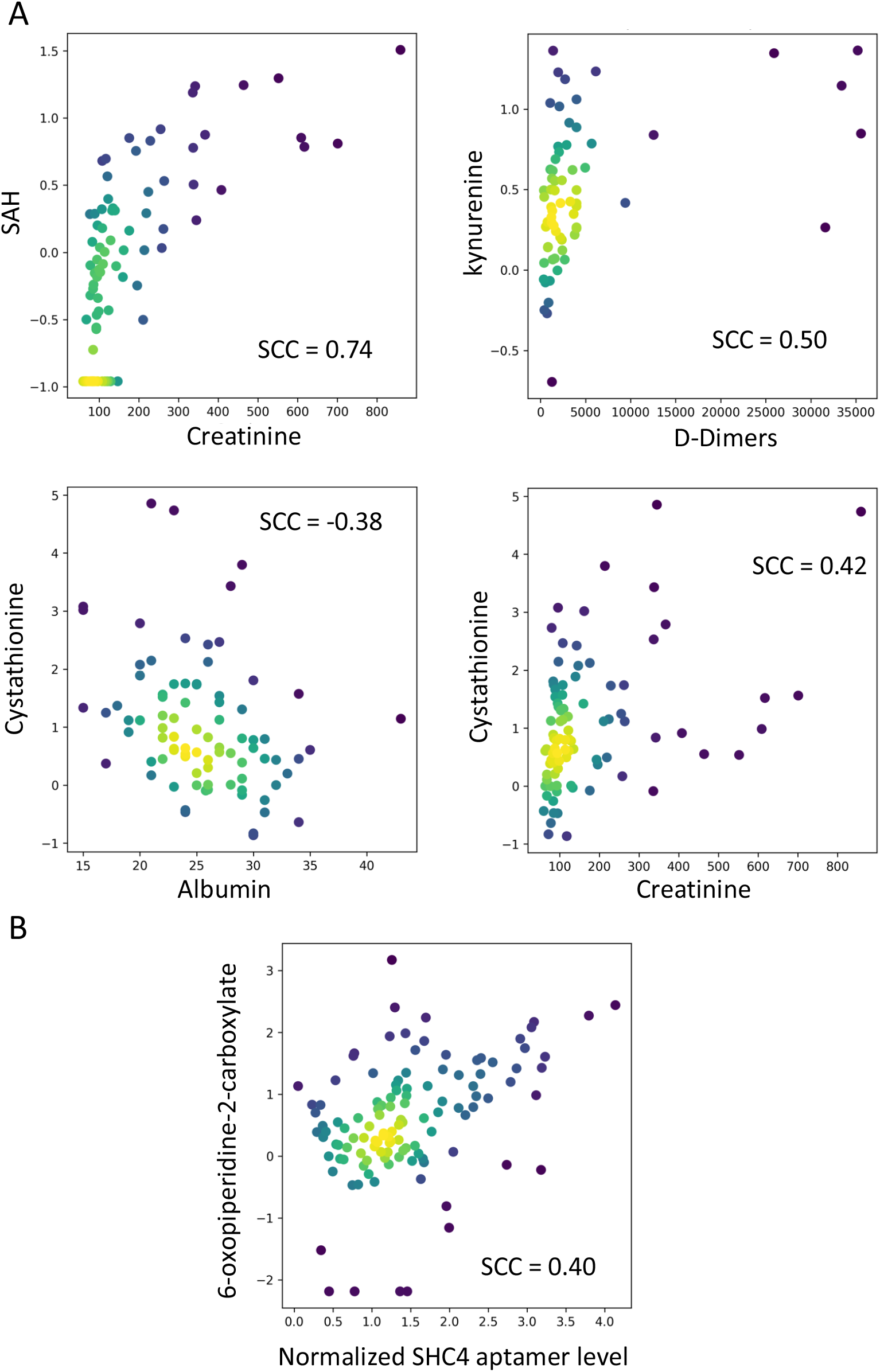
Correlation between different variables in EP6. Each circle in the scatter plots correspond to a patient in EP6. The circles are colored based on the density of points surrounding them for clearer visualization. SCC corresponds to Spearman’s rank correlation coefficient. A) Scatter plots reflecting the correlation between metabolites (y-axis) and blood variables (x-axis). B) Scatter plot corresponding to correlation of 6-oxopiperidine-2-carboxylate and SHC4.

**Figure 5:**
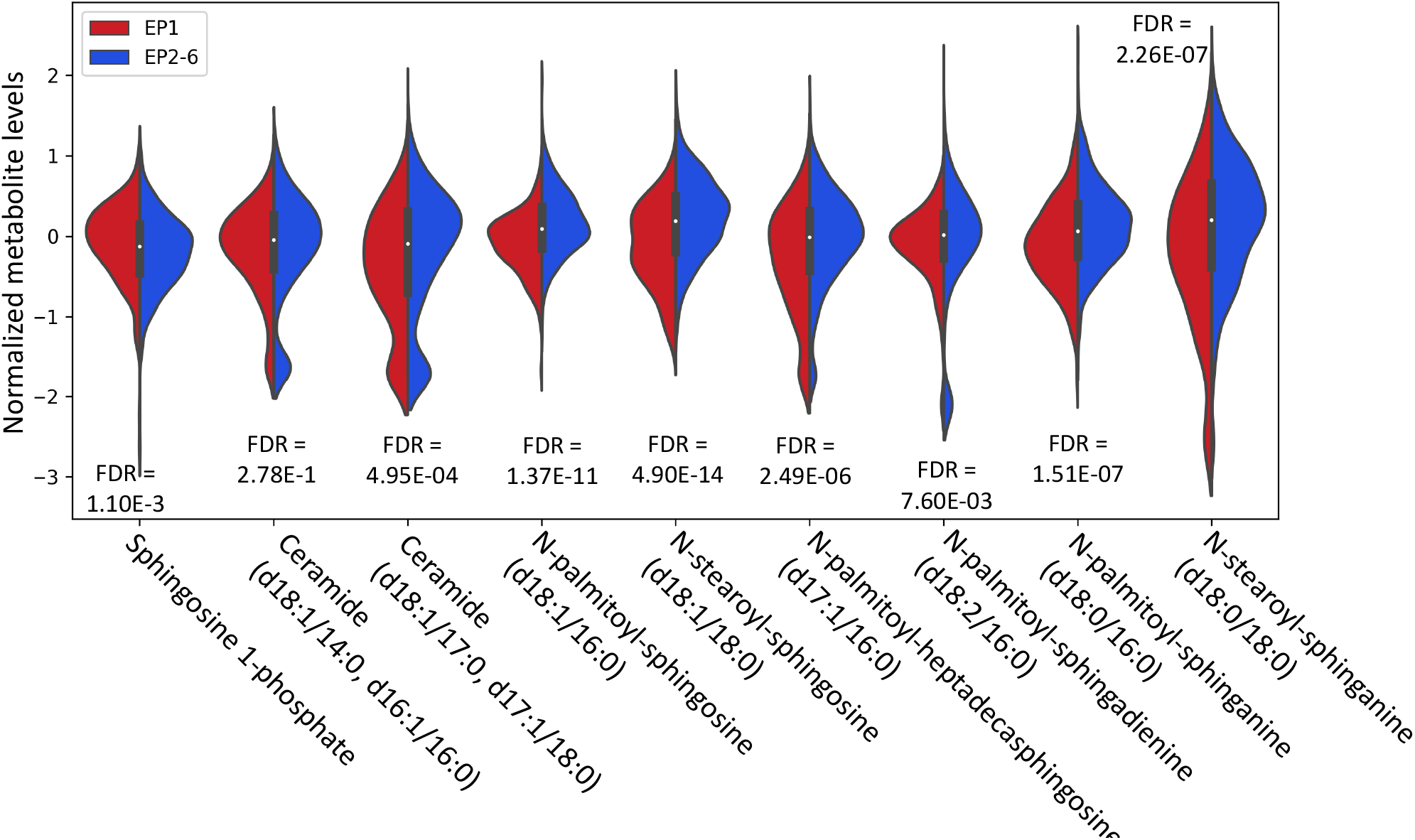
Distribution of metabolite levels in EP1 versus other EPs. The violin plots show the distribution of metabolite levels for patients in EP1 (red) versus EP2-6 (blue). Two-sided Mann–Whitney U (MWU) tests followed by Benjamini–Hochberg method for adjusting the p-values were used to calculate false discovery rates.

### Low levels of sphingosine-1 phosphate are associated with COVID-19 ARDS

Strikingly, sphingosine-1 phosphate was enriched in EP1 and depleted in EP6 (Figures 3 and 5), confirming the previously reported negative association of this metabolite with COVID-19 ARDS (18). Accordingly, the aptamer detecting neutral ceramidase, an enzyme converting ceramides into sphingosine, is enriched in EP1 (FDR = 2.65E-10, Table S1). While changes in protein abundance as detected by aptamers may not necessarily reflect changes in enzymatic activity, it is interesting to note that dihydroceramides and ceramides are depleted in EP1 (Figure 5, Table S1). Conversely, dihydroceramides and ceramides are significantly enriched in EP6 (Figure 3, Table S1). Moreover, one of the top aptamers found enriched in EP6 (FDR = 2.10E-56, Figure 1) is serine palmitoyltransferase 2 (SPTLC2), an enzyme that is responsible for the rate-limiting step in *de novo* sphingolipid biosynthesis (19, 20).

### Mechanical ventilation in EP6 is associated with lipoprotein metabolism

As noted in the introduction, mechanical ventilation in COVID-19 is associated with a high mortality rate. Within EP6, 55 out of the 118 individuals received mechanical ventilation. Within all EP6 members that share the overall similar aptamer signature, we identified a subset of aptamers and metabolites that were significantly different between those with or without mechanical ventilation (Figure 6A and Table S4). The second most enriched aptamer (Table 2 and S4), angiopoietin-like 3 (ANGPTL3), corresponds to a liver-secreted regulator of lipoprotein metabolism (21), which may reflect alterations observed in fatty acid metabolites (Table S4). There were no significant differences between the two groups in terms of sex, age, or body mass index (BMI) (Figure 6B-6C and Table S4). Finally, we investigated whether molecular information (proteomic and/or metabolomic) was associated with mechanical ventilation duration. We identified alpha-L-iduronidase (IDUA) to be inversely correlated with duration of ventilation in the patients with mechanical ventilation in EP6 (Spearman’s rank correlation = −0.60, FDR = 2.57E-2, Figure 6D and Table S4). Overall, these results point to an association between altered lipoprotein metabolism and mechanical ventilation in EP6.

**Table 2:** Aptamers associated with mechanical ventilation in EP6. In this table, p-values are obtained using a two-sided Mann–Whitney U test and is corrected for multiple tests using Benjamini–Hochberg FDR. A value of +1 (−1) in the “sign” column shows that the value of aptamer was higher (lower) in the samples of 55 patients that received mechanical ventilation in EP6, compared to the rest of EP6 members. Only 14 targets with FDR < 1.2E-02 are shown. The full list is provided in Table S4.

| Target | Target Full Name | p-values | FDR | Direction |
| --- | --- | --- | --- | --- |
| GGE2D | G antigen 2D | 2.93E-08 | 1.46E-04 | +1 |
| ANGPTL3 | Angiopoietin-related protein 3 | 2.76E-06 | 4.58E-03 | +1 |
| Luteinizing hormone | Luteinizing hormone | 2.55E-06 | 4.58E-03 | -1 |
| CRBS | Beta-crystallin S | 1.01E-05 | 8.37E-03 | -1 |
| NCAM-120 | Neural cell adhesion molecule 1; 120 kDa isoform | 8.78E-06 | 8.37E-03 | -1 |
| SIA10 | Type 2 lactosamine alpha-2;3-sialyltransferase | 7.36E-06 | 8.37E-03 | -1 |
| LRP1 | Prolow-density lipoprotein receptor-related protein 1 | 2.77E-05 | 1.16E-02 | -1 |
| CLM2 | CMRF35-like molecule 2 | 2.70E-05 | 1.16E-02 | -1 |
| THS7A | Thrombospondin type-1 domain-containing protein 7A | 1.65E-05 | 1.16E-02 | -1 |
| LAP | Cytosol aminopeptidase | 3.04E-05 | 1.16E-02 | -1 |
| CXCL16 | C-X-C motif chemokine 16 | 2.64E-05 | 1.16E-02 | +1 |
| FSH | Follicle stimulating hormone | 2.52E-05 | 1.16E-02 | -1 |
| VEGF sR2 | Vascular endothelial growth factor receptor 2 | 3.27E-05 | 1.16E-02 | -1 |
| ASGR1 | Asialoglycoprotein receptor 1 | 2.97E-05 | 1.16E-02 | +1 |

**Figure 6:**
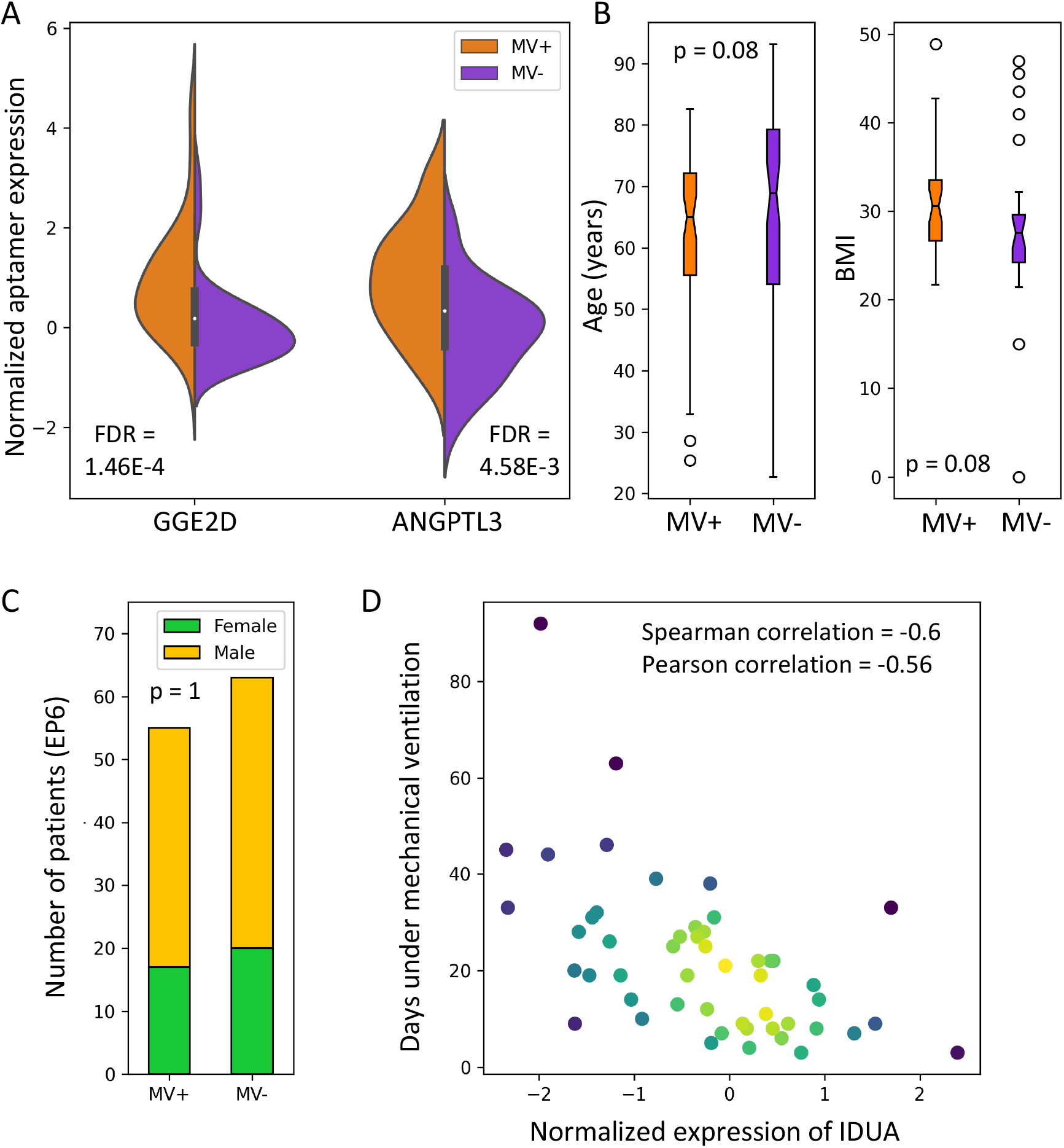
Mechanical ventilation in EP6. A) The violin plots show the distribution of aptamer expressions for patients in EP6 that received mechanical ventilation (MV+) versus those that did not (MV-). Two-sided Mann–Whitney U (MWU) tests followed by Benjamini–Hochberg method for adjusting p-values were used to calculate false discovery rates. B) Distribution of age and BMI of MV+ and MV-patients in EP6. P-values are calculated using two-sided MWU tests. C) The status of sex for MV+ and MV-patients in EP6. P-value is calculated using a Fisher’s exact test. D) The scatter plot shows the correlation between expression of IDUA aptamer and number of days that patients received mechanical ventilation (n = 50).

### SHC4 genotype and protein expression levels are associated with higher odds of belonging to EP6

To obtain independent support for the involvement of specific proteins identified using aptamers, we leveraged genome-wide association study (GWAS) genotyping data corresponding to these patients. We identified 25 single nucleotide variations (SNVs) distributed in 13 annotated genes that were below a p-value threshold of 1E-4 and distinguished EP6 from the rest (Table 3). We then investigated each of the SNVs to which we could assign a gene and an aptamer, to assess whether their protein product in circulation was differentially regulated by the genotype (Table 4). We discovered two genes, *SHC4* (encoding SHC adaptor protein 4) and *CACNA2D3* (encoding calcium voltage-gated channel auxiliary subunit alpha2 delta3) for which there was a significant association between genotype and protein expression levels (p-values < 0.05). While *CACNA2D3* may have mild impact on EP6 membership (odds ratio = 0.61, Table 4), SHC4 was one of the top-enriched aptamers (position 32 out of 4,985, Figure 1) for inclusion in EP6, with an odds ratio of 11.98 for the protein product and 2.00 for the alternate allele, respectively. Finally, to gain further insight into the potential role of SHC4 in COVID-19 disease severity, we investigated the metabolites associated with SHC4 in EP6. The only significantly correlated metabolite was 6-oxopiperidine-2-carboxylate (Spearman’s rank correlation = 0.40, FDR = 0.012, Figures 4, S1-S2), where the p-values were corrected for the number of metabolites using BH FDR. Therefore, the GWAS analysis revealed that the signaling adaptor protein SHC4 may play an important mechanistic role in contributing to severe disease pathology.

**Table 3:** SNVs differentiating EP6 against all other endophenotype clusters.

| SNV ID | Position | Gene Symbol | SNV <sup>1</sup> | MAF | HWE | OR <sup>2</sup> | p-value |
| --- | --- | --- | --- | --- | --- | --- | --- |
| rs1394671 | chr5:7348567 | - | G > A | 0.240 | 0.095 | 2.28 | 2.41E-06 |
| rs12186698 | chr5:168880668 | *SLIT3* | T > C | 0.087 | 0.247 | 2.93 | 3.72E-06 |
| rs11625406 | chr14:50049474 | - | C > A | 0.438 | 0.137 | 0.47 | 1.03E-05 |
| rs11862889 | chr16:83828886 | - | T > C | 0.104 | 0.446 | 2.62 | 1.07E-05 |
| rs2995918 | chr4:37905775 | *TBC1D1* | C > T | 0.494 | 0.776 | 0.49 | 1.20E-05 |
| rs16897810 | chr5:68415296 | - | G > A | 0.093 | 0.001 | 2.67 | 1.43E-05 |
| rs2376263 | chr17:35436659 | *SLFN13* | G > A | 0.261 | 0.073 | 2.01 | 2.48E-05 |
| rs4790712 | chr17:1614620 | *SLC43A2* | G > A | 0.441 | 0.003 | 1.97 | 2.50E-05 |
| rs2294566 | chr20:41472939 | *CHD6* | C > A | 0.239 | 0.265 | 2.04 | 2.78E-05 |
| rs7164451 | chr15:48921859 | *SHC4* | G > A | 0.386 | 0.053 | 1.89 | 3.01E-05 |
| rs57664621 | chr8:22808443 | *PEBP4* | G > C | 0.174 | 0.016 | 2.16 | 3.07E-05 |
| rs28482919 | chr3:14903094 | *FGD5* | T > C | 0.187 | 0.053 | 0.32 | 3.20E-05 |
| rs6559283 | chr9:89712560 | - | T > C | 0.375 | <0.001 | 1.96 | 3.27E-05 |
| rs657075 | chr5:132094425 | - | A > G | 0.104 | 0.328 | 2.49 | 4.28E-05 |
| rs56235109 | chr15:62424001 | *TLN2* | A > G | 0.235 | 0.366 | 0.38 | 4.67E-05 |
| rs7620057 | chr3:179473377 | *GNB4* | T > C | 0.099 | 1.000 | 2.51 | 4.67E-05 |
| rs10466868 | chr12:131455375 | - | T > G | 0.108 | 0.401 | 2.51 | 5.94E-05 |
| rs2236798 | chr1:18735127 | *PAX7* | A > G | 0.054 | 0.114 | 2.90 | 6.11E-05 |
| rs3774814 | chr4:5464702 | *STK32B* | C > G | 0.205 | <0.001 | 0.36 | 6.75E-05 |
| rs4497815 | chr19:22903215 | - | G > A | 0.216 | 0.205 | 2.07 | 7.52E-05 |
| rs6765694 | chr3:54601810 | *CACNA2D3* | G > A | 0.404 | 0.833 | 0.51 | 7.66E-05 |
| rs2797773 | chr6:37559045 | - | C > T | 0.422 | 0.000 | 0.53 | 8.06E-05 |
| rs17014760 | chr4:129419725 | - | A > A | 0.316 | 0.000 | 1.89 | 9.33E-05 |
| rs10948260 | chr6:45835559 | - | G > A | 0.366 | 0.189 | 1.81 | 9.52E-05 |
| rs12035677 | chr1:232391209 | - | A > G | 0.063 | 0.000 | 3.02 | 9.95E-05 |

Abbreviations used: SNV = Single nucleotide variation, HWE = Hardy Weinberg Equilibrium, MAF = Minor allele frequency, OR = Odd ratio.
<sup>1</sup> SNV are described following GWAS annotations: reference allele > alternative allele (e.g., G > A).
<sup>2</sup> Logistic regression analyses using additive model adjusted for the 2 principal components.

**Table 4:** Association between genotypes and aptamer expression levels

| SNV | Gene Symbol | Nearest gene | Aptamers normalized expression <sup>1</sup> |  |  |  | Multiple logistic regression analyses <sup>3</sup> |  |  |  |
| --- | --- | --- | --- | --- | --- | --- | --- | --- | --- | --- |
|  |  |  | HM<sub>ref</sub> | HTZ | HM<sub>alt</sub> | p-val <sup>2</sup> | Aptamers OR | p-val | SNV OR | p-val |
| rs6765694 | *CACNA2D3* | - | -0.09<br>(±0.89) | 0.01<br>(±1.10) | 0.24 *<br>(±0.99) | **0.011** | 0.61<br>(0.46-0.79) | 2.5E-4 | 0.52<br>(0.37-0.74) | 2.4E-4 |
| rs10948260 | - | *CLIC5* | 0.02<br>(±0.92) | 0.02<br>(±1.09) | -0.02<br>(±0.97) | 0.796 | 0.68<br>(0.55-0.83) | 1.7E-4 | 1.83<br>(1.35-2.48) | 8.8E-5 |
| rs657075 | - | *IL3* | -0.01<br>(±1.01) | 0.05<br>(±0.99) | 0.32<br>(±0.46) | 0.328 | 1.37<br>(1.14-11.64) | 8.2E-4 | 2.49<br>(1.60-3.88) | 5.2E-5 |
| rs7164451 | *SHC4* | - | -0.07<br>(±0.90) | 0.01<br>(±1.05) | 0.23 *<br>(±1.16) | **0.017** | 11.98<br>(7.61-18.87) | 8.4E-27 | 2.00<br>(1.30-3.09) | 1.8E-3 |
| rs12186698 | *SLIT3* | - | 0.00<br>(±1.02) | 0.07<br>(±1.06) | 0.15<br>(±0.89) | 0.509 | 0.82<br>(0.64-1.06) | 0.135 | 2.99<br>(1.89-4.73) | 2.7E-6 |
| rs56235109 | *TLN2* | - | -0.03<br>(±0.98) | 0.06<br>(±1.03) | 0.04<br>(±1.27) | 0.353 | 0.58<br>(0.45-0.75) | 2.7E-5 | 0.38<br>(0.23-0.60) | 4.7E-5 |

Abbreviations used: SNV = Single nucleotide variation, HM<sub>ref</sub> = Homozygotes for the reference allele, HM<sub>alt</sub> = Homozygotes for the alternative allele, HTZ = Heterozygotes, OR = Odd ratio.
<sup>1</sup> Aptamers' normalized levels of expression are reported as mean (± standard deviation). Normalization steps for aptamer expressions are described in Methods.
<sup>2</sup> p-value, standard ANOVA analyses followed by Tukey *post hoc* analyses. Asterisks (\*) identify difference between HM<sub>ref</sub> and HM<sub>alt</sub> genotypes.
<sup>3</sup> Multiple logistic regression analyses models include aptamers expression values, SNV genotypes (additive model) and the two principal components. OR are reported with 95% confidence intervals in parentheses.

## Discussion

In this study, we leveraged the molecular features associated with endophenotypes discovered using unsupervised hierarchical clustering of circulating proteome of SARS-CoV-2 positive hospitalized participants to BQC19 to improve our understanding of pathobiology. This approach enabled the identification of the following molecular features: 1) the level of Sphingosine-1 phosphate is highest in EP1 and lowest in EP6, reflecting an inverse association with time to admission to ICU and/or death; 2) interleukins and FGFR signaling are molecular pathways associated with severe COVID-19; 3) genetic and environmental determinants of SHC4 expression are associated with inclusion in EP6; 4) the combined proteomic and metabolomic signature of EP6 supports a pathologic role for neutrophil-associated procoagulant activity in COVID-19 ARDS; and 5) mechanical ventilation in EP6 is associated with altered lipoprotein metabolism.

### COVID-19 ARDS molecular pathology: a counterbalance between ceramides and sphingosine is associated with critical illness

The datasets used in this study carry rich molecular information on mechanisms of disease for COVID-19. EP6 is characterized by its enrichment in ARDS (50% vs 9% in EP1), acute kidney injury (50%) and liver dysfunction (22%) (9). Low levels of Sphingosine 1-phosphate (S1P) are associated with ARDS and have been shown to be associated with greater ICU admission and decreased survival in COVID-19 (18), and is supported by this study where S1P levels were found depleted in EP6 but enriched in EP1 (the endophenotype with the best prognostic). The study also supports a shunting away from sphingosine towards more pro-inflammatory ceramides that are associated with metabolic disorders (22), suggesting a counterbalance between ceramides and sphingosine, where the former is associated with poorer outcomes during critical illness, whereas higher levels of the latter is associated with more favorable outcomes in ARDS.

### Molecular mechanisms of mechanical ventilation in EP6: a potential role for ANGPTL3

ANGPTL3 is a liver-secreted enzyme that can inhibit lipoprotein and endothelial lipase (23). Loss-of-function of ANGPTL3 is associated with hypolipidemia (24). Therefore, increased circulating levels of ANGPTL3 would be expected to raise plasma levels of triglycerides and phospholipids which is consistent with the observed metabolomic profiles (Table S4). Whether these alterations are sufficient to increase the likelihood of mechanical ventilation remains speculative. Nevertheless, it is interesting to note that administration of medium- and long-chain triglycerides can aggravate gas exchange abnormalities in ARDS (25) and may warrant the preclinical investigation of ANGTPL3 antagonists like evinacumab or other functionally similar therapeutic agents (26) in ameliorating outcomes in ARDS. As for the duration of mechanical ventilation, we discovered an inverse correlation with IDUA, a lysosomal enzyme required for the degradation of heparan sulfate and dermatan sulfate (27). Our current data cannot distinguish between the presence of this enzyme as a biomarker or as an active mediator that influences the course of COVID-19-associated acute respiratory failure.

### Multimodal profiling of EP6 supports a role for neutrophil-derived procoagulant activity-mediated organ damage in COVID-19

COVID-19 ARDS has been found to be more frequently associated with thrombotic complications than non-COVID-19 ARDS (28, 29). In addition, histopathologic evidence of pulmonary microthrombi is frequently observed in autopsies of individuals deceased with COVID-19 (30). Early reports suggested a role for neutrophil-mediated release of NETs contributing to endothelial dysfunction as a mechanism of microthrombosis in COVID-19 ARDS (31-42). The neutrophil-derived serprocidins cathepsin G, neutrophil elastase, and proteinase-3, are significantly enriched in EP6. In addition to their roles in bacterial killing, serprocidins couple innate immunity to coagulation, where they promote coagulation by enhancing tissue factor and factor XII-dependent coagulation (43). Serprocidins are also important regulators of NETosis (44). In pediatric sepsis, another NETosis-associated protein olfactomedin 4 (OLFM4) (10), enriched in EP6, is associated with increased odds of having greater organ failure and death (45). The procoagulant environment associated with COVID-19-associated organ damage is further supported by the alteration in metabolic profiles in EP6. Phosphatidinylethanolamines (PE) become exposed at the surface of cell membranes upon exposure to stress, inflammation, and cell death (46, 47). In a Syrian hamster model, infection with SARS-CoV-2 markedly increased circulating PE expression in the animals that were fed a high salt, high fat diet, demonstrating the interaction between infection and metabolic disorder (48). Furthermore, the membranes of azurophilic granules, which contain serprocidins, are enriched in PE (49). Accordingly, EP6 is associated with decreased platelet counts, increased D-Dimers, international normalized ratio (INR) and activated partial thromboplastin time (aPTT) (9), hallmarks of abnormally activated coagulation pathways that are associated with serious and often lethal complication of sepsis (50). Accordingly, we report an association between kynurenine and D-dimer, pointing to an involvement of tryptophan metabolism in abnormal coagulation. Interestingly, TNFSF18, a T-cell receptor that promotes leukocyte adhesion to endothelial cells (51), is enriched in EP6. A SNV found in the proximity of TNFSF18 was found associated with severity of COVID19 in the western Indian population (52). Unfortunately, this SNV was absent from the array used in BQC19 preventing the confirmation of that observation, but it is nevertheless suggestive of a link between TNFSF18, kynurenine, and abnormal coagulation in severe COVID-19. Collectively, these findings point to immuno-thrombosis activity in EP6 in response to SARS-CoV-2 infection that is associated with abnormal coagulation. Recent results from the Antithrombotic Therapy to Ameliorate Complications of COVID-19 (ATTACC) study showed anticoagulation to be efficacious in moderate but not severe COVID-19 disease (53, 54), revealing a need for alternative management approaches in critically ill patients.

### Novel molecular markers of COVID-19 pathology: FGFR and SHC4

Two novel molecular factors associated with COVID-19 ARDS that were identified by our study are FGFR and SHC4. Circulating levels of the pro-angiogenic FGF-2 has been associated with COVID-19 severity and creatinine levels in a study of 208 SARS-CoV-2 positive participants (55). It is also noteworthy that the use of nintedanib, an inhibitor of FGFR, vascular endothelial growth factor receptor (VEGFR), and platelet-derived growth factor receptor (PDGF-R) that is approved for use in interstitial lung disease, improved pulmonary inflammation and helped three middle-aged obese COVID-19 patients with markedly impaired lung function wean off mechanical ventilation (56). While the adaptor protein SHC4 has not been experimentally demonstrated to modulate FGFR signaling, a 12-gene biomarker signature associated with melanoma contains FGFR2, FGFR3 and SHC4 (57). It is attractive to speculate that SHC4 may act downstream of FGFR or other associated growth factor receptors, favoring heightened procoagulant activity associated with COVID-19 ARDS. In view of the limited knowledge of this understudied member of the SHC family, we looked at the metabolites associated with SHC4 to gain insights into its possible functions. Interestingly, 6-oxopiperidine-2-carboxylate (the only metabolite with a significant correlation with SHC4 in EP6) was found to be negatively associated with glomerular filtration rate in a GWAS study of kidney disease and hypertension in African Americans (58). Acute kidney injury is a frequent complication of acute liver failure (59), and liver dysfunction is associated with abnormal coagulation (60). Thus, the overall molecular information coming from the multi-modal analysis of EP6 points to a triad between liver function, kidney function, and hemostasis that becomes dysfunctional following ARDS-associated inflammation driven by SARS-CoV-2 infection. Whether the presence of SHC4 in circulation is a marker of dysfunction of this triad, or an active mediator, remains to be determined. Moreover, the identity of the cells expressing SHC4 leading to its presence in the circulation is not known. Taken together, our identification of FGFR and SHC4 signaling pathways distinguishing EP6 from other endophenotypes provides a rationale for investigation of their potential utility as biomarkers of severe disease activity and/or the use of antagonists of those pathways to treat severe COVID-19 disease manifestations.

## Methods

### Datasets and preprocessing

We obtained data corresponding to clinical, genomics, metabolomics, and proteomics (circulating proteome) of n = 731 hospitalized and SARS-CoV-2 positive patients (based on qRT-PCR) from the Biobanque Québécoise de la COVID-19 (BQC19; www.quebeccovidbiobank.ca). The circulating proteome measurements obtained using a multiplex SOMAmer affinity array (SomaLogic, 4,985 aptamers) between April 2020 and April 2021 were preprocessed and used to identify six endophenotypes (EP1-EP6). The details of the cohort, circulating proteome data processing, and identification of endophenotypes are provided in the accompanying manuscript (9).

In this study, we used the batch-normalized, missing-values-imputed, and log-transformed version of the BQC19 metabolomic data (1,435 metabolites). BQC19 GWAS imputation data were generated by Tomoko Nakanishi at Brent Richards lab, Jewish General Hospital and McGill University. Detailed codes used for generating the data can be found in: https://github.com/richardslab/BQC19_genotype_pipeline.

### Analyses of the GWAS dataset

For the GWAS analyses, annotation of SNVs were done using the biomaRt package (61) from R (62), and all analyses were done using R version 4.1.3. Quality control steps were derived in majority from a 2017 QC tutorial article (63). At the beginning, we had 867,450 markers and 2,429 samples. We imported Plink format data into R using the “read_plink” function from genio R package (64). We removed 103,592 non ACGT bi-allelic markers. We calculated the predicted sex by looking at the rate of homozygote markers on chromosome 23. We removed 3,588 markers with call rates < 98%, 448,932 monomorphic markers and markers with MAF < 0.05, and 28,092 markers with Hardy–Weinberg equilibrium < 1E-6 (calculated by the “HWE.exact” function from the genetics package (65) from R. For the EP6 cluster group analyses, we additionally removed 1,747 samples that were not in the cluster analysis, 8 samples with a sex discrepancy (based on predicted sex calculated earlier and reported sex), and 3 samples with a heterozygosity rate > 3 standard deviations. We finally removed a pair of samples which had approximately the same genome, likely due to an error of manipulation. As we could not know which one was correct, we removed both. We also found 2 pairs of individuals with a pi-hat of ∼0.5 (meaning first degree relatives), so we kept the one sample per pair with the higher call rate. All other pairs of individuals had a pi-hat < 0.21 that is judged acceptable considering our population. Pi-hats were calculated with the “snpgdsIBDMoM” function from SNPRelate R package (66). At the end of quality controls, 283,246 markers on 655 samples had been used to perform association analyses.

To perform the principal component analyses (PCA), we took a subsample of independent markers (pruning) with a maximum sliding window of 500,000 base pairs and a linkage disequilibrium (LD) threshold of 0.2 using the “snpgdsLDpruning” function from the SNPRelate R package. We ran the PCA with the “snpgdsPCA” function from SNPRelate package from R. The first 2 principal components (PCs) were considered significant.

For the GWAS analyses of the 283,246 remaining markers between EP6 cluster compared to all others, we used a logistic regression model, with the dichotomous variable indicating if the participants belong to the EP6 cluster as the outcome variable. We used the additive model for markers as an independent variable, and we adjusted the models with the first two PCs. Odds ratio and p-values were calculated on each model. Quantile–quantile (Q–Q) plots have been performed as quality control of the models; p-values were plotted using “qqplot.pvalues” function from gaston R package (67) (data not shown). We compared the aptamers’ normalized level of expression between the three groups of genotypes for each studied gene by performing standard analysis of variance (ANOVA) analyses followed by Tukey *post hoc* tests (referred to in this study as pQTL analysis). Since aptamers tested are limited compared to SNVs, we fixed significance p-value threshold below 1E-4 to report more SNVs instead of the more common 1E-5 suggestive threshold. Finally, to identify if EP6 may be characterized by the aptamers and the SNVs, we performed multiple logistic regression analysis including aptamers expression values, SNV genotypes (additive model), and the two principal components. Odds ratios are reported with 95% confidence intervals.

### Metabolomic pathway characterization of EP6

The 1,435 metabolites measured were organized into 122 sub-pathways in the original dataset (denoted as “SUB_PATHWAYS”). We first identified metabolites whose values were significantly higher or lower in EP6 compared to other EPs (two-sided MWU test, FDR<0.01). Then, we used these metabolites to perform pathway enrichment analysis (one-sided Fisher’s exact test) based on 122 pathways. The resulting p-values were then corrected for multiple tests using Benjamini– Hochberg FDR.

### Statistics

Several non-parametric tests, including the MWU test, Fisher’s exact test, and Spearman’s rank correlation, were used in this study. Benjamini–Hochberg FDR was used to adjust the p-values for multiple tests.

### Study approval

The study was approved by the Institutional Ethics Review Board of the “Centre intégré universitaire de santé et de services sociaux du Saguenay-Lac-Saint-Jean” (CIUSSS-SLSJ) affiliated to the Université de Sherbrooke [protocol #2021-369, 2021-014 CMDO – COVID19].

## Supporting information

STROBE Checklist

## Data Availability

Input data corresponding to the cohort can be obtained form BQC19 (www.quebeccovidbiobank.ca). The data generated as a result of the analyses are provided as tables and supplementary tables.

## Acknowledgements

This work was made possible through open sharing of data and samples from the Biobanque Québécoise de la COVID-19, funded by the Fonds de recherche du Québec - Santé, Génome Québec, the Public Health Agency of Canada and, as of March 2022, the Ministère de la Santé et des Services Sociaux du Québec. We thank all participants to BQC19 for their contribution. This study was supported by the Fonds de recherche du Québec - Santé (FRQS)-Cardiometabolic Health, Diabetes and Obesity Research Network (CMDO)-Initiative. This work was also supported by Natural Sciences and Engineering Research Council of Canada (NSERC) grant RGPIN-2019-04460 (AE).

