## Supplementary File S1 for "Molecular pathology of acute respiratory distress syndrome, mechanical ventilation and abnormal coagulation in severe COVID-19"

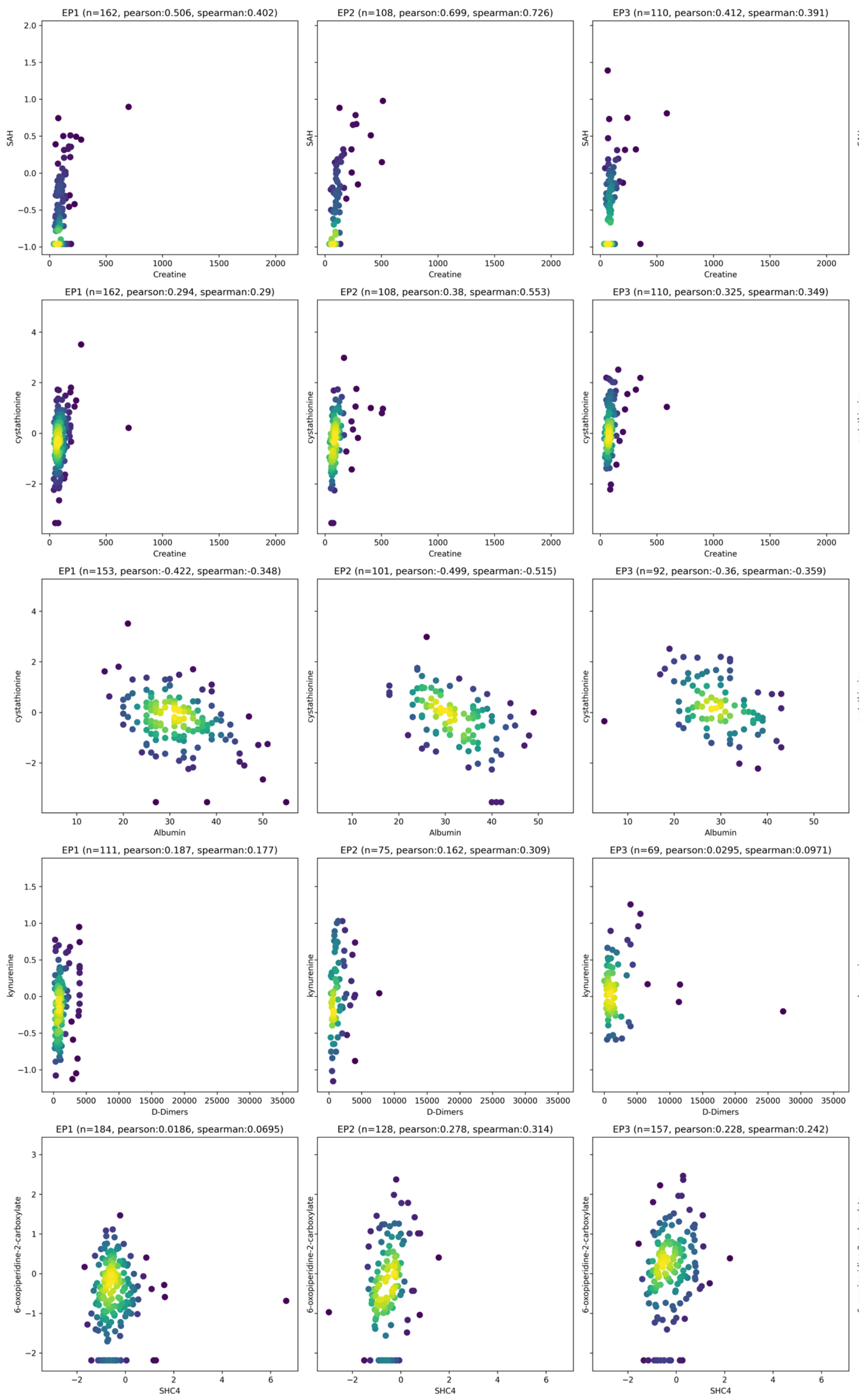


**Figure S1:** Correlation between different variables in EP1-EP3.
Each circle in the scatter plots corresponds to a patient in an EP. Each column of panels corresponds to a different EP (EP1 to EP3, from left to right).

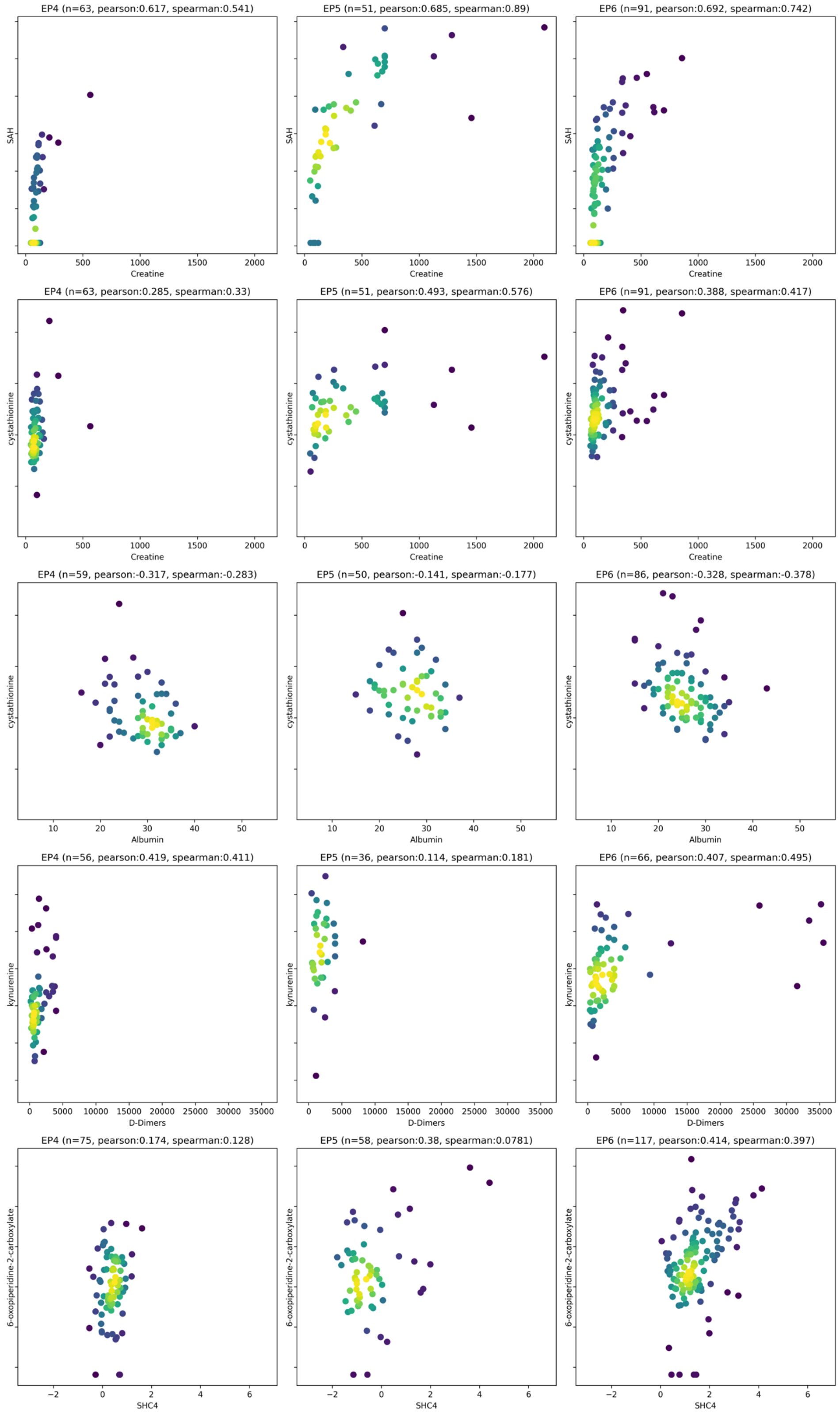


**Figure S2:** Correlation between different variables in EP4-EP6.

Each circle in the scatter plots corresponds to a patient in an EP. Each column of panels corresponds to a different EP (EP4 to EP6, from left to right).

**Supplementary Tables**

**Table S1:** Aptamer enrichment and pathway analysis. The table is provided as a separate excel file.

**Table S2:** Metabolite enrichment and sub-pathway analysis for each endophenotype. The table is provided as a separate excel file.

**Table S3:** Correlation between blood variables and aptamers in endophenotype 6 (EP6). The table is provided as a separate excel file.

**Table S4:** Mechanical ventilation characteristics of patients in endophenotype 6 (EP6). The table is provided as a separate excel file.
